# Prediction of donor heart acceptance for transplant and its clinical implications – results from the Donor Heart Study

**DOI:** 10.1101/2023.10.30.23297809

**Authors:** Brian Wayda, Yingjie Weng, Shiqi Zhang, Helen Luikart, Thomas Pearson, R. Patrick Wood, Javier Nieto, Bruce Nicely, PJ Geraghty, John Belcher, John Nguyen, Nikole Neidlinger, Tahnee Groat, Darren Malinoski, Jonathan Zaroff, Kiran K. Khush

**Affiliations:** Division of Cardiovascular Medicine, Department of Medicine, Stanford University School of Medicine, Stanford, CA; Quantitative Sciences Unit, Department of Medicine, Stanford University School of Medicine, Stanford, CA; LifeLink Foundation, Norcross, GA; LifeGift Organ Procurement Organization, Houston, TX; Gift of Life Michigan, Ann Arbor, MI; Donor Network of Arizona, Tempe, AZ; New England Donor Services, Waltham, MA; Division of Transplant Surgery, Department of Surgery, University of California San Francisco, San Francisco, CA; Department of Surgery, University of Wisconsin School of Medicine and Public Health, Madison, WI; Department of Surgery, Division of Trauma, Critical Care, and Acute Care Surgery, Oregon Health and Science University, Portland, OR; Division of Research, Kaiser Permanente Northern California, Oakland, CA

**Keywords:** heart transplantation, organ procurement, machine learning, models, decision-support, donor selection

## Abstract

**Background:** Despite a shortage of potential donors for heart transplant in the United States (US), most potential donor hearts are discarded. We evaluated predictors of donor heart acceptance in the US and applied modern analytic methods to improve prediction.

**Methods:** We included a *nationwide* (2005 – 2020) cohort of potential heart donors in the US (n = 73,948) from the Scientific Registry of Transplant Recipients and a more recent (2015 – 2020) rigorously phenotyped cohort of potential donors from the Donor Heart Study (*DHS*; n = 4,130). We identified predictors of acceptance for heart transplant in both cohorts using multivariate logistic regression, incorporating time-interaction terms to characterize their varying effects over time. We fit models predicting acceptance for transplant in a 50% training subset of the DHS using multiple machine learning algorithms and compared their performance in the remaining 50% (test) subset.

**Results:** Predictors of donor heart acceptance were similar in the *nationwide* and *DHS* cohorts. Among these, older age has become increasingly predictive of discard over time while other factors – including those related to drug use, infection, and mild cardiac diagnostic abnormalities - have become less influential. A random forest model (area under the curve 0.90, accuracy 0.82) outperformed other prediction algorithms in the test subset and was used as the basis of a novel web-based prediction tool.

**Conclusions:** Predictors of donor heart acceptance for transplantation have changed significantly over the last two decades, likely reflecting evolving evidence regarding their impact on post-transplant outcomes. Real-time prediction of donor heart acceptance, using our web-based tool, may improve efficiency during donor management and heart allocation.

**Clinical Perspective:** Predictors of donor heart acceptance for transplantation have changed significantly over the last two decades. Donor age has become increasingly influential while several other factors have become less so - likely reflecting the lack of evidence regarding their impact on post-transplant outcomes. Our web-based tool can enable real-time prediction of donor heart acceptance, and thereby improve efficiency during donor management and heart allocation.

## Introduction

New heart transplant (HT) listings in the United States (US) (4,588 in 2020) have consistently outpaced the number of transplants performed (3,715 in 2020).^1^ As a result, many candidates wait several months or longer for transplant and hundreds die annually while waiting. Despite this unmet demand, the majority of potential donor hearts are not used for transplant.^2^ Common reasons for non-use include older donor age, left ventricular (LV) dysfunction or hypertrophy, cardiovascular comorbidities, or other risk factors.^3^

Given these consequences of the donor organ scarcity, high discard rates are justified only if they “select out” donor risk factors with an adverse impact on post-transplant outcomes. For example, a recent meta-analysis identified a reliable association between donor age and recipient mortality.^4^ For other putative risk factors listed above, such an association is plausible but the evidence is less robust.^5^ Naturally, donor heart discard practices should evolve in concert with the evidence base. To judge whether they have requires careful examination of the factors that drive donor heart acceptance (vs. discard) and how they have evolved over time.

Critical to the goal of rational donor heart selection is the role played by organ procurement organizations (OPOs), who evaluate and manage potential donors in the hours before their organs are offered for transplant or discarded.^6^ OPOs aim to maximize the utilization of viable organs for transplant - an objective that requires complex decisions involving multiple (sometimes competing) priorities. For example, pursuing coronary angiography and serial echocardiograms to evaluate a “high-risk” donor heart can delay the recovery of other solid organs and potentially compromise their quality.^7^ Accordingly, it may be best to defer such intensive cardiac evaluation when the likelihood of yielding a viable donor heart is very low; such a decision is difficult in real-time, and hinges upon the basic question: how likely is this donor to be accepted for heart transplant?

In this study, we assess current predictors of donor heart acceptance in the US, how they have changed over the last two decades, and whether they reflect evolving evidence on which donor characteristics impact post-transplant outcomes. Second, we apply these predictors using modern machine learning methods to enable real-time prediction of donor heart acceptance.

## Methods

### Data sources

Our primary data source was the Donor Heart Study (DHS), an observational prospective cohort study of potential heart donors enrolled from February 2015 to May 2020, which has been described previously.^8^ The DHS was coordinated at Stanford University and conducted at 8 OPOs across the US (listed in **Table 1**). The DHS was approved by the Stanford University Institutional Review Board (Protocol 31461) and by the research oversight committee at each participating OPO.

**Table 1:**
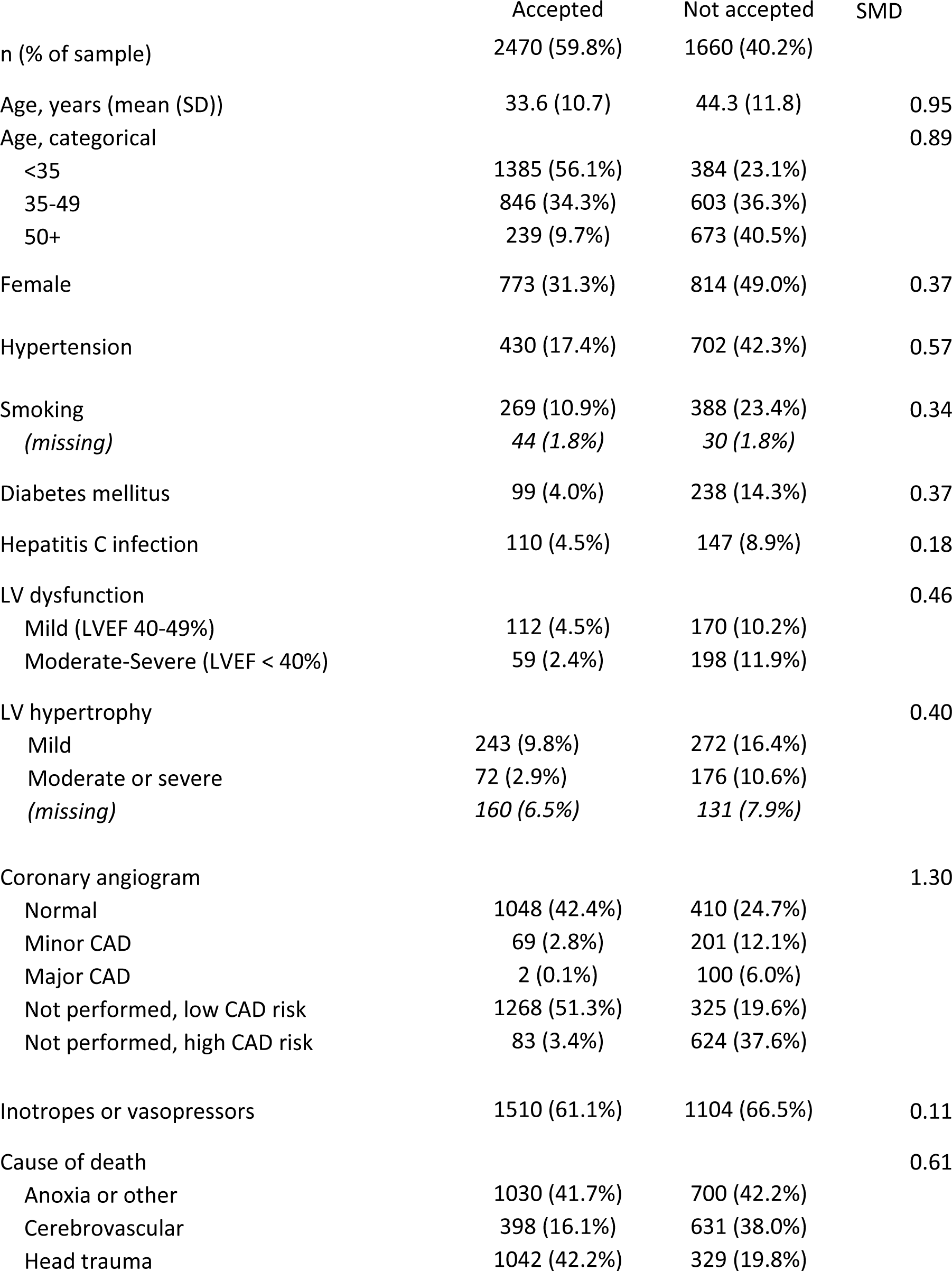
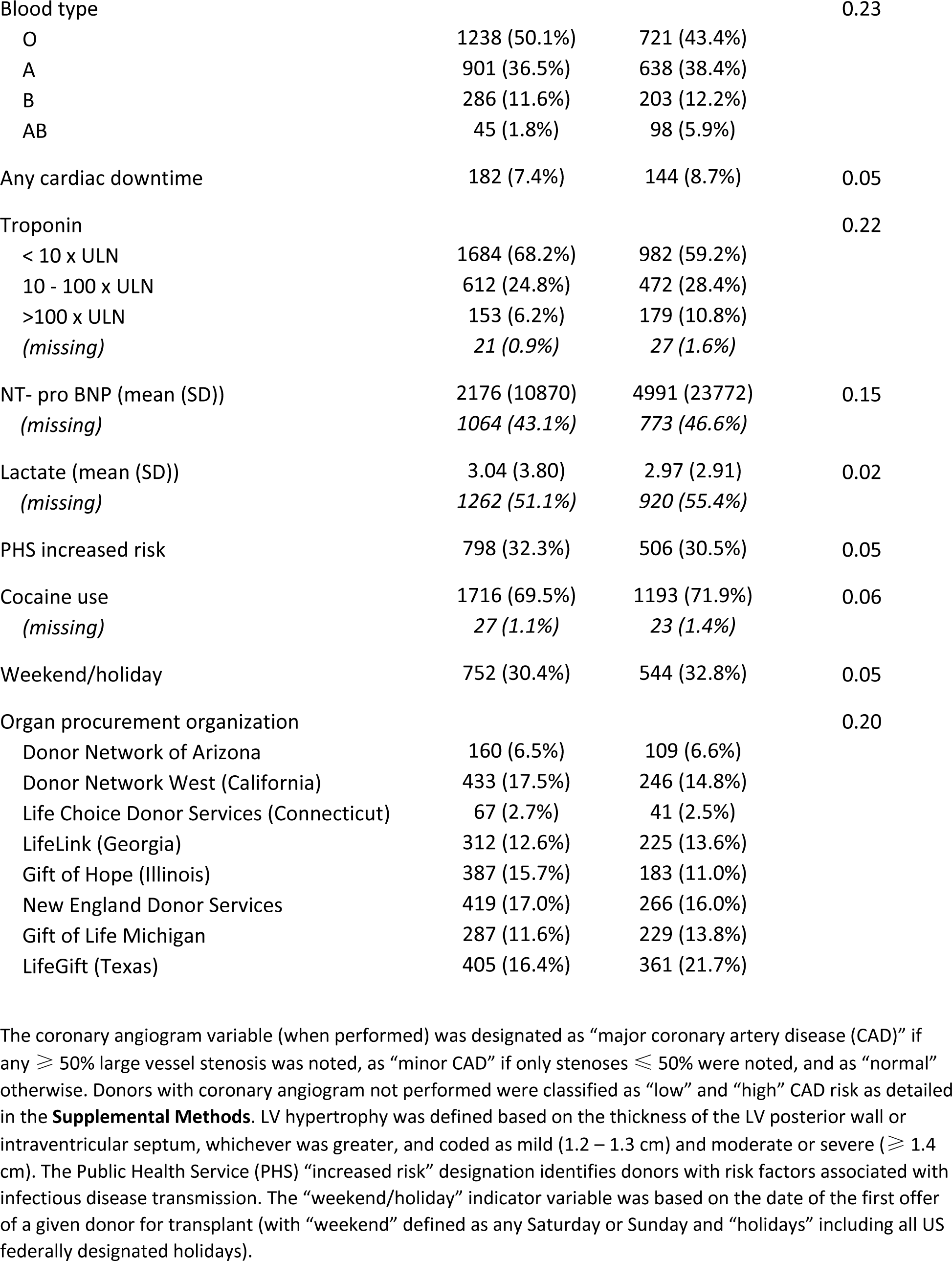
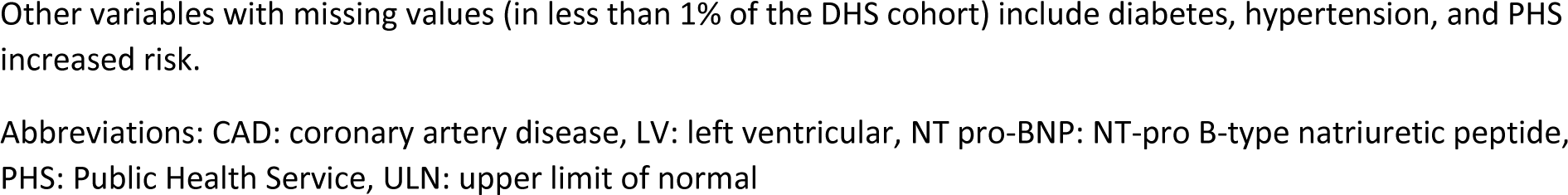
Descriptive characteristics of Donor Heart Study donors, by acceptance for heart transplantation.

We also used data from the Scientific Registry of Transplant Recipients (SRTR), which includes a record of all donors in the US from whom at least one solid organ was recovered for transplant.^1^ The data reported here has been supplied by the Minneapolis Medical Research Foundation as the contractor for SRTR. The interpretation and reporting of these data are the responsibility of the authors and in no way should be seen as an official policy of or interpretation by the SRTR or the United States government.

These two data sources offered complimentary advantages. SRTR’s larger sample size and duration was necessary to characterize the how predictors of donor heart acceptance have changed over time (as detailed below in “Inference models”). The DHS characterized contemporary donors in greater detail, allowing more robust prediction of acceptance (as detailed below in “Prediction models”).

### Variables

Our outcome variable was *donor acceptance for HT*. Covariates (listed in **Table 1**) included donor demographics, comorbidities, and diagnostic findings. We focused on those felt to be intrinsic to the donor and excluded those that often reflect clinical management decisions (e.g. blood pressure, electrolyte levels). For covariates measured at multiple timepoints, we utilized only those obtained at the beginning of donor management, as this is the time at which prediction of donor heart acceptance would have most clinical utility. Further details on the outcome variable and selected covariates are included in the **Supplemental Methods**.

### Cohort definitions

The full *DHS cohort* was divided randomly into equally sized derivation (training) and validation (test) subsets. We defined a *nationwide* cohort (including both DHS and non-DHS OPOs) which included all potential heart donors in the US over a longer period (February 2005 – May 2020).

In each of these cohorts, we included donors aged 18-65 years and excluded any with declaration of circulatory death (DCD), a history of myocardial infarction, or the absence of a recorded left ventricular ejection fraction (LVEF). The rationale for these exclusion criteria was to limit each cohort to donors who were at least considered for heart transplant (as opposed to other solid organ transplant only). The construction of each cohort is illustrated with a CONSORT diagram (**Supplemental Figure 1**).

### Statistical analysis

#### a. Descriptive analysis

For each cohort, we reported mean and standard deviation (SD) for continuous variables and frequencies and proportions for categorical variables. To compare donors by outcome status, we reported standardized mean difference (SMD), a comparative measure of effect size between groups. Consistent with prior guidelines, the magnitude of effect was considered “small” if SMD = 0.2 – 0.5, “moderate” if SMD = 0.5 – 0.8, and “large” if SMD > 0.8.^9,10^

#### b. Inference models

Multivariable logistic regression models were fit separately in both the DHS and nationwide cohorts to identify independent associations of each covariate with acceptance for transplant. We fit an additional logistic regression model in the nationwide cohort in which calendar year and interaction terms between calendar year and each covariate were added; the purpose of this model was to assess for changes in the influence of each covariate over time. We used p < 0.05 as the threshold for statistical significance for both “stand-alone” and time-interaction terms in these inference models. We did not apply corrections for multiple comparisons given the models were evaluated for descriptive purposes instead of for hypothesis testing.

In each of these inference models, we accounted for missing donor covariates using multiple imputation, with 10 imputed datasets. Lactate and NT-pro B-type natriuretic peptide (NT-pro BNP) were log-transformed prior to inclusion in logistic regression models. Other covariates were represented as categorical variables as detailed in **Table 1**.

#### c. Prediction models

To develop our prediction model, we compared the performance of multiple machine learning algorithms including logistic regression, random forest and least absolute shrinkage and selection operator (LASSO).^11^ An R-shiny^12^ web-based application was created for our final prediction model.

Using the DHS derivation cohort, we constructed prediction models using each of the three algorithms and the full set of covariates detailed above. The performance of each was assessed in the DHS validation cohort using model accuracy and area under the curve (AUC). We computed 95% confidence intervals (CIs) of these AUCs using bootstrapping methods. Calibration plots were also produced to evaluate the discrimination of the algorithms. In each of these prediction models, we accounted for missing donor covariates using multiple imputation, with five imputed datasets.

We excluded coronary artery disease (CAD) as a covariate in all prediction models. Our rationale was that the extent of CAD is often unknown at the start of donor management, when a predictive model would have the most utility; coronary angiography is typically performed later and may be deferred if donor heart acceptance is deemed unlikely.

## Results

### Donor characteristics, by cohort and outcome status

Descriptive characteristics of donors in the DHS cohort, including the 59.8% (n = 2470) accepted for transplant and the 40.2% (n = 1660) not accepted for transplant, are shown in **Table 1**. There were large differences between these two subgroups in donor age and coronary angiogram findings and small or moderate differences in several other risk factors. Specifically, accepted donors had mean age of 33.6 years (vs. 44.3 for non-accepted donors) and lower prevalence of CAD, smoking, diabetes mellitus, hypertension, cerebrovascular cause of death, LV dysfunction (i.e. LVEF < 50%), and LV hypertrophy.

Similar differences in donor characteristics by acceptance status were found in the nationwide cohort (n = 73,948), as detailed in **Supplemental Table 1**. However, the proportion accepted for transplant (45.4%; p < 0.001) was significantly lower in the nationwide (vs. DHS) cohort.

### Predictors of acceptance and their change over time

Associations between donor characteristics and acceptance for transplant after multivariable adjustment are shown in **Figure 1**. In both the DHS and nationwide long-term cohorts, factors most strongly associated with non-acceptance for transplant (odds ratio < 0.4 for all) included older age, hepatitis C infection, LV dysfunction, CAD, and blood type AB.

**Figure 1:**
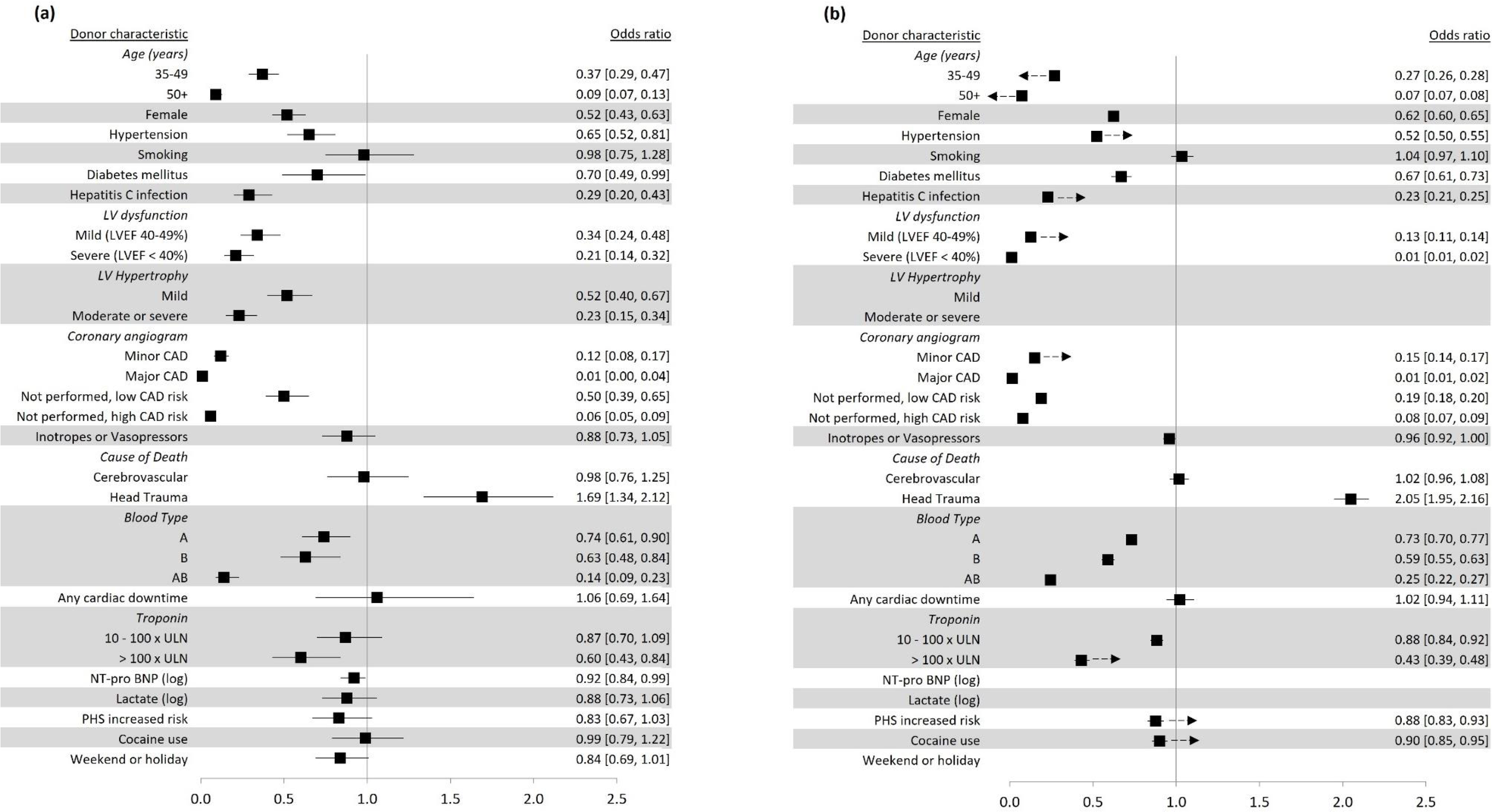
Forest plots of adjusted associations between donor characteristics and acceptance for HT in (a) DHS and (b) nationwide cohorts and their change over time. Shown are odds ratios and 95% Cis (in brackets) from multivariable logistic regression models as detailed in **Methods**. Referent groups include Age < 35 years, normal coronary angiogram, normal LV function (LVEF ≥ 50%), male sex, anoxia or other cause of death, ABO type O, absence of LV hypertrophy, and troponin < 10 x ULN. Arrows (panel b) indicate the direction of all statistically significant time interactions. Abbreviations: CAD: coronary artery disease, DHS: Donor Heart Study, HT: heart transplantation, LV: left ventricular, NT pro-BNP: NT-pro B-type natriuretic peptide, PHS: Public Health Service

In both cohorts, smaller (odds ratios 0.4 to 0.8) but significant associations with non-acceptance were observed for diabetes mellitus, female sex, blood types A and B, hypertension, and troponin > 100 times the upper limit of normal. Head trauma as cause of death was associated with a significantly higher likelihood of acceptance in both the DHS (OR: 1.69 [1.34 – 2.12]) and nationwide long-term (OR: 2.05 [1.95 – 2.16]) cohorts.

The absence of a coronary angiogram was significantly associated with non-acceptance, although with varying effect size by cohort and presence of CAD risk factor(s). LV hypertrophy (measured only in the DHS cohort) was a significant predictor of non-acceptance, both when mild (OR: 0.52 [0.40 – 0.67]) and when moderate or severe (OR: 0.23 [0.15 – 0.34]). Other covariates measured only in the DHS cohort – including NT-pro BNP (OR: 0.92 [0.84 – 0.99]), lactate (OR: 0.88 [0.73 – 1.06]), and weekend/holiday offers (OR: 0.84 [0.69 – 1.01]) predicted non-acceptance with varying statistical significance.

In a model employing time interaction terms in the nationwide long-term cohort (as detailed in the **Methods**), several predictors of non-acceptance had decreasing influence over time (had time-interaction terms that were positive and statistically significant). These are indicated by arrows in **Figure 1b** and include: Hepatitis C positivity, mild LV dysfunction (LVEF 40-49%), minor CAD, hypertension, elevated troponin (> 100 times upper limit of normal), cocaine use, and Public Health Service “increased risk” (which captures drug use and other high-risk behaviors, as detailed in the **Supplemental Methods**). In contrast, both age 35-49 years and age ≥ 50 years were increasingly predictive of non-acceptance over time.

### Prediction model: comparison, validation, and web-based application

A detailed comparison of performance characteristics between the random forest, logistic regression, and LASSO-based prediction models is shown in **Supplemental Table 2**. In summary, the random forest model outperformed the other algorithms by all metrics, with an accuracy of 0.82 and AUC of 0.90 in the validation sets. Its receiver operator characteristic curve and calibration plot are shown in **Figure 2**. By comparison, the logistic and LASSO models had an AUC of 0.83 in both derivation and validation sets.

**Figure 2:**
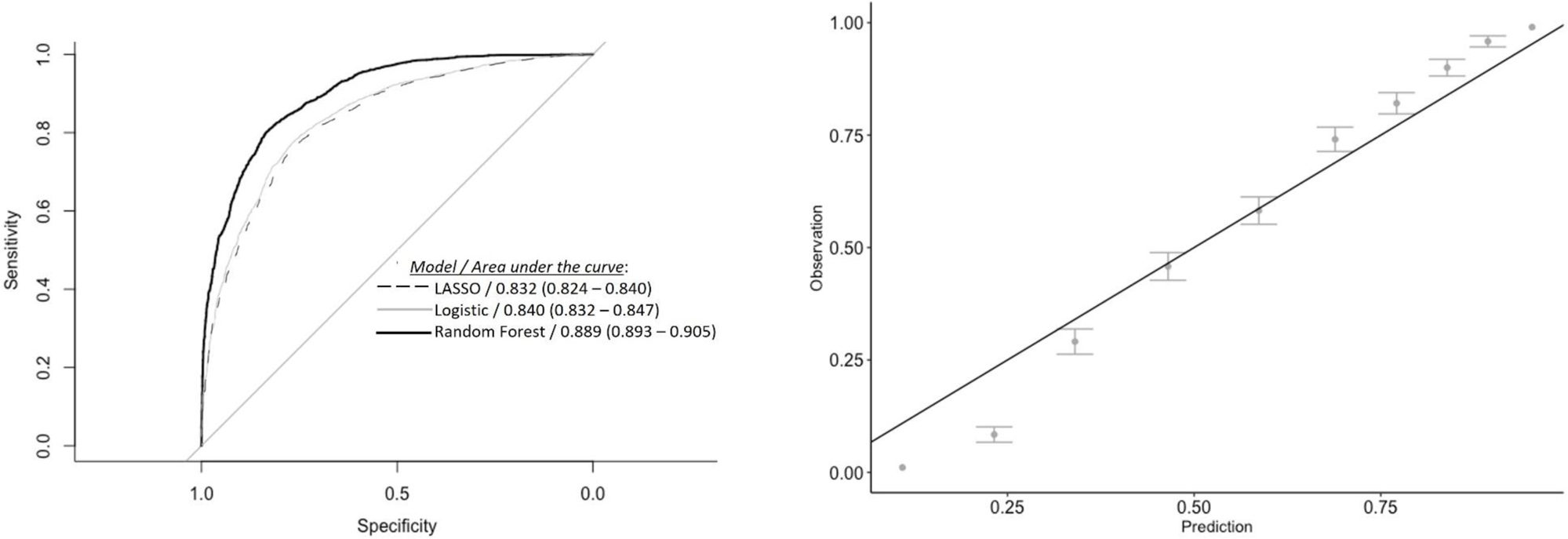
Receiver operating characteristic curves (a) and calibration plot (b; Random Forest model only) for prediction models in test dataset.

Our best-performing model (Random Forest) was used as the basis for our web-based prediction tool (ToP-HAT: Tool to Predict Heart Acceptance for Transplant), available at https://qsushiny.shinyapps.io/TOPHAT/ and depicted in **Figure 3**. It includes 18 donor characteristics as inputs, with the relative importance^13^ of each shown in **Supplemental Figure 2**.

**Figure 3:**
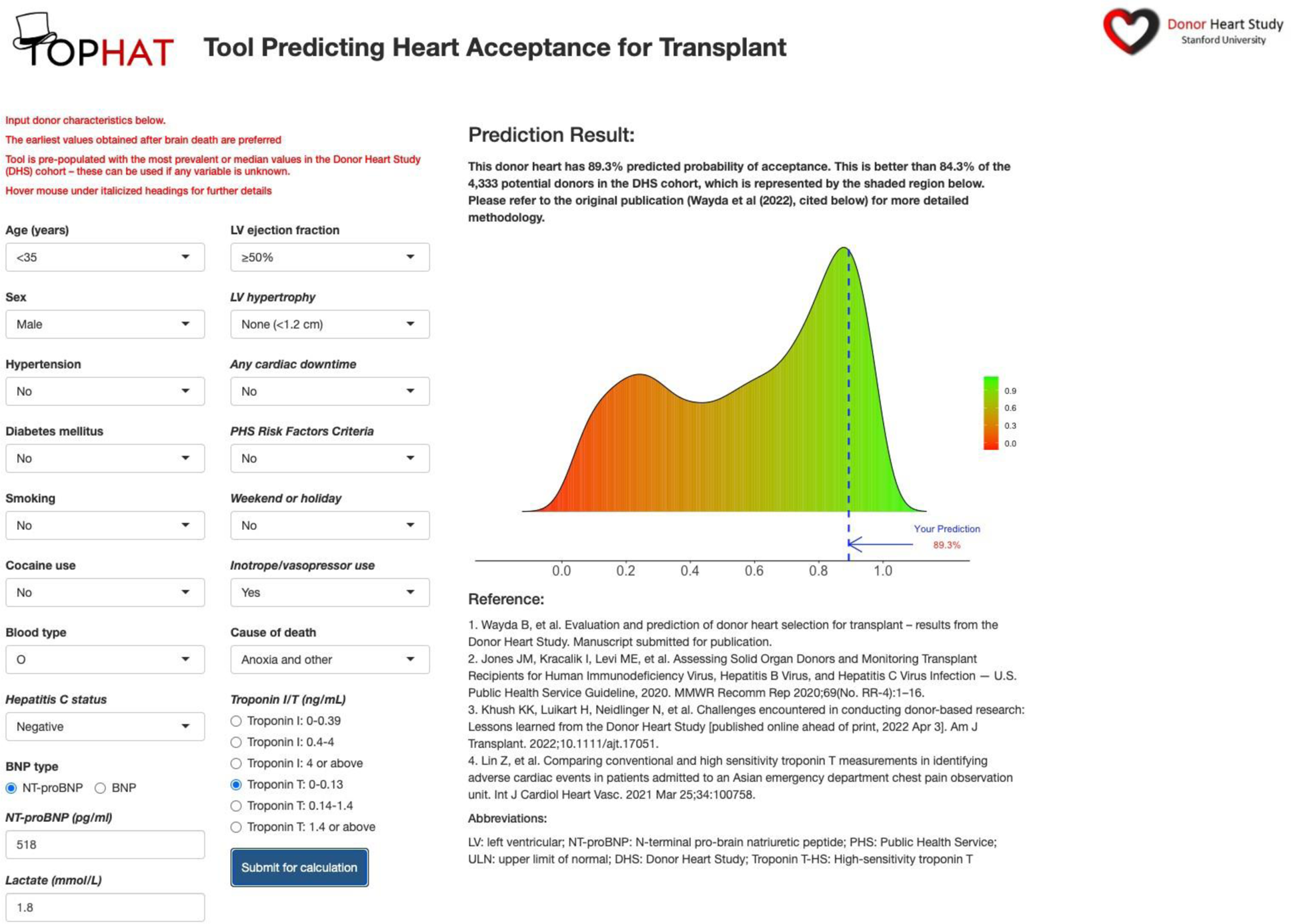
Screenshot of web-based tool predicting heart acceptance for transplant (ToP-HAT)

## Discussion

Using the Donor Heart Study prospective cohort of over 4,000 carefully phenotyped potential heart donors, our study evaluated the donor characteristics that determine heart acceptance for transplant. As expected, abnormalities of the potential donor heart itself are most influential, but a wider array of other non-cardiac donor characteristics independently predict acceptance (vs. discard). We also find that these predictors of donor heart acceptance have changed significantly over the last two decades. Together, our findings offer insights into contemporary donor heart selection practices, which warrant scrutiny amid a persistent donor organ shortage.

As a practical extension of these analyses, we present a Tool to Predict Heart Acceptance for Transplant (ToP-HAT) – a prediction model based on modern machine learning methods that performed well in a validation cohort.

### Evaluation of donor heart selection practices

Any scrutiny of donor heart selection practices must acknowledge the following: there is no scientific consensus regarding which factors should be considered when evaluating a potential donor for heart transplant. Guidelines on heart donor selection (detailed in **Supplemental Table 3)** have become less specific over time, granting greater deference to subjective clinical judgement. ^6,14,15^ The most recent of these (from 2020) focuses on donor age and cardiac diagnostic findings as important to consider, but omits the array of non-cardiac characteristics cited in prior iterations – a trend reflecting the lack of rigorous evidence regarding their effects on post-HT outcomes.^16^

We find that contemporary donor selection mirrors this guidance; age ≥ 50 years, severe LV dysfunction (LVEF < 40%), moderate or severe LVH, and major CAD were the strongest predictors of non-acceptance. Meanwhile, those “risk factors” related to drug use – which have no independent association with post-HT outcomes^17–20^ and have been dropped from the guidelines - have had a diminishing influence on donor heart selection over time.

While agreement between guidelines and practice is always encouraging, exactly *how much* – and *at what threshold* – older age should deter donor acceptance is unclear. That donor age > 50 confers a ten-fold reduction in the odds of acceptance – even after adjusting for age-associated risk factors – seems excessive in light of the following: 1) individual centers report excellent outcomes using carefully-selected donors over 50 years (i.e. those without accompanying risk factors),^21–24^ 2) 50+ year-old donors are used routinely in other countries (e.g. ∼20% of all HTs in the Eurotransplant consortium),^25^ and 3) the “age effect” appears to be mediated by the presence of other risk factors (e.g. CAD and prolonged ischemic time).^26,27^

The profound influence of cardiac diagnostic abnormalities – even in their common and milder forms - warrants similar scrutiny. While the harm posed by severe CAD or severe LV hypertrophy may be self-evident, the impact of a minor (<50%) stenosis (OR for acceptance: 0.12) or mild hypertrophy (OR: 0.52) on post-HT outcomes is unclear. Donor LV dysfunction also reduces the odds of acceptance by three- to five-fold – despite prior evidence from the DHS and elsewhere that it is usually reversible and not associated with post-HT outcomes.^28–30^ The dire consequences of the ongoing donor heart scarcity demand further research into *which* (not whether) donors with these milder and/or transient abnormalities can safely be used for HT.

Clinical intuition suggests that any adverse effect of donor hypertension or diabetes on post-HT outcomes would be mediated by CAD or LV hypertrophy. Yet both remain associated with non-acceptance even after multivariate adjustment; by implication, a donor with a completely normal echocardiogram and coronary angiogram will still be “penalized” by the presence of these comorbidities. Their relevance to donor selection should be further investigated and perhaps explicitly addressed in consensus guidelines.

Our analysis is the first to evaluate the association of donor BNP with acceptance for HT. We find its influence is small but significant (OR 0.92; p = 0.038) - perhaps appropriately so, given the virtual absence of data on how to interpret donor BNP levels. Yet we recently found that it predicts the reversibility of donor LV dysfunction;^30^ accordingly, the broader prognostic utility of donor BNP and other acute biomarkers warrants further study.

### Prediction of donor heart acceptance

We present a novel prediction model to assess the likelihood of donor heart acceptance. Our model is available for real-world application, in the form of the web-based ToP-HAT calculator. Similar models already exist - including one using logistic regression that is provided online by SRTR^31^ – but we find that use of a random forest algorithm offers improved prediction. The superiority of modern machine learning methods has been noted in other HT-related contexts^32,33,34^

We envision the real-time use of ToP-HAT by OPO teams, as they evaluate and manage potential donors prior to offering organs for transplant. For context, some potential donors are promptly disqualified from HT due to known heart disease (often as the cause of death); at the other extreme are those with no cardiovascular comorbidities and a reassuring echocardiogram. In between these extremes is a large pool of heart donor candidates whose viability for HT is not immediately clear, perhaps due to risk factors for CAD and/or an abnormal screening echocardiogram. As detailed in **Figure 4**, whether to pursue (or defer) time-consuming and expensive cardiac evaluation for these donors is a weighty decision – and could be guided by use of ToP-HAT.

**Figure 4:**
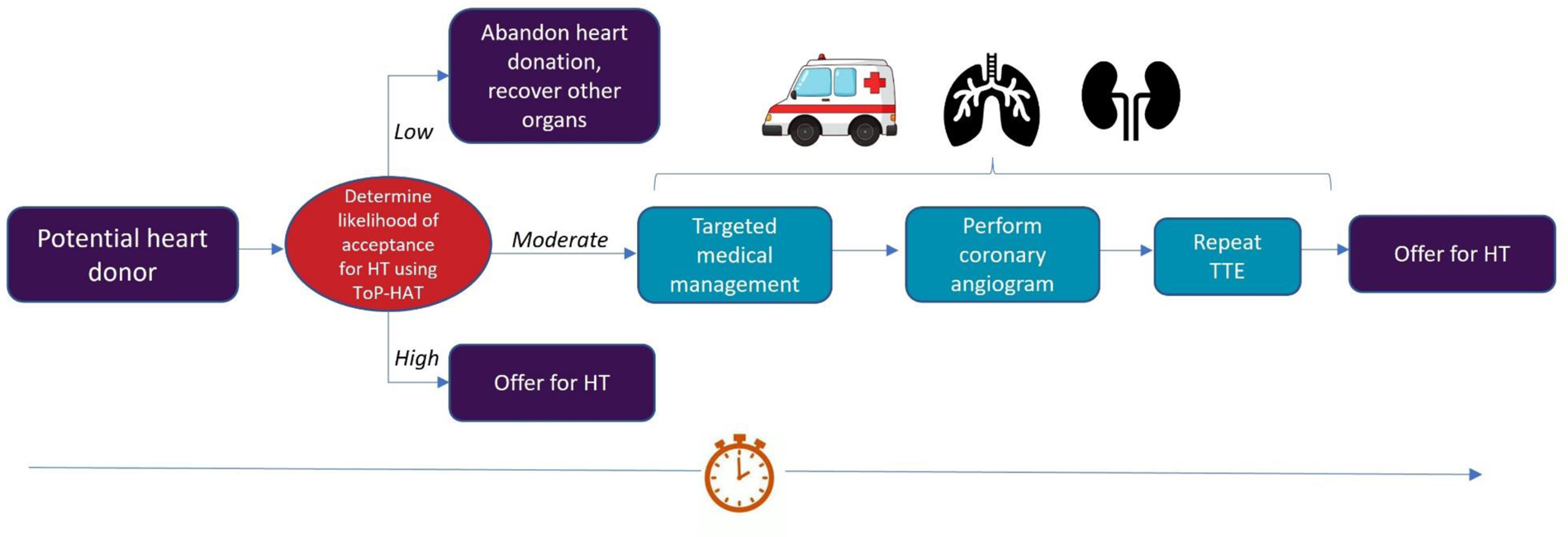
A potential scenario for implementation of ToP-HAT during donor evaluation. For many potential donors, their viability for use in HT is not clear at the early stages of donor management. Making this determination can require extensive cardiac workup including serial echocardiograms and/or a coronary angiogram; depending on local availability, the latter may require transfer to a different center. The resulting delay in organ recovery is resource-intensive, including augmented inotropic therapy and administration of thyroid hormone or corticosteroids, and can compromise the quality of other solid organs, particularly when the donor is hemodynamically unstable. The probability of acceptance for HT – estimated early on using ToP-HAT – can help guide the decision to pursue further cardiac workup. When this probability fails to meet some reasonable threshold, then deferring evaluation for potential HT may be warranted. Abbreviations: HT: heart transplantation, ToP-HAT: Tool Predicting Heart Acceptance for Transplantation, TTE: transthoracic echocardiogram

Our model can also be used by transplant clinicians and centers to supplement their own subjective donor assessment. Those on “donor call”, evaluating a specific offer might wonder: *would other centers be likely to accept this donor?* Centers with low donor utilization could ask, *are we turning down donors that other centers would elect to accept*? ToP-HAT can be used to answer both of these questions.

### Study limitations

Given our use of a large, geographically diverse derivation cohort, we suspect that our model’s component predictors and overall performance will translate to non-DHS OPOs. Unfortunately, external validation of our model was not feasible, as there is no large donor cohort data of comparable detail.

As our study shows, donor acceptance practices are in flux – likely influenced by evolving evidence, by the size and composition of the potential donor (and recipient) pool, and by a myriad of other time- and place-specific factors. Our model does not capture this variation; its predictions reflect “aggregate” behavior and must be interpreted in the local context. Its use to predict acceptance for DCD donors (excluded from our cohort) is not recommended.

Despite its unprecedented detail, the DHS omits some potentially relevant donor datapoints (e.g. invasive hemodynamic measures (which are not systematically collected during donor management) and specific circumstances of death) that could influence acceptance for HT. Their omission constrains the performance of our prediction model and our identified predictors of acceptance may be partly capturing the influence of these unobserved variables.

In conclusion, our study has 1) evaluated the wide array of factors that determine donor acceptance for HT and 2) developed a model to enable real-time prediction of donor heart acceptance. Both can help improve the efficiency of the organ evaluation and allocation process, and can inform clinical practice and policy efforts to safely increase donor heart utilization for HT, thereby granting more patients access to this life-saving therapy.

## Data Availability

No new data?? were created during this study

## List of Supplemental Items

Detailed Methods

Supplemental Table 1: Descriptive characteristics of potential heart donors in the US nationwide cohort (2005 – 2020), by outcome status

Supplemental Table 2: Detailed comparison of model performance in logistic regression, LASSO, and Random Forest prediction models

Supplemental Table 3: Historical summary of expert guidelines regarding donor heart selection criteria

Supplemental Figure 1: Study CONSORT diagram for (a) DHS and (b) nationwide cohorts

Supplemental Figure 2: Relative importance of donor covariates in Random Forest prediction model

